# Knowing when, what, and how to push: Examining the operationalization of task challenge in routine stroke rehabilitation practice using focused video analysis

**DOI:** 10.64898/2026.09.28.26364226

**Authors:** Emeline Gomes, Gemma Alder, Felicity A.S. Bright, Nada Signal

**Affiliations:** School of Allied Health, Auckland University of Technology, Auckland, New Zealand; Person Centred Rehabilitation Research Centre, Auckland University of Technology, Auckland, New Zealand

**Keywords:** Stroke rehabilitation, Challenge, Video analysis, Therapeutic relationship, Person-centered care, Dosage, Intensity

## Abstract

**Background:** Task challenge is recognized as a core principle of neurorehabilitation and is frequently mentioned within stroke rehabilitation guidelines. However, guidance for operationalizing challenge in clinical practice remains limited. Examining how therapists and people with stroke currently use challenge in practice may help address this gap.

**Objective:** To examine how challenge is operationalized in routine stroke rehabilitation practice.

**Methods:** This study used focused video analysis to examine eight video-recorded stroke rehabilitation sessions across physiotherapy, occupational therapy, and speech-language therapy contexts. Video data were supplemented by observational memos. Analysis was conducted iteratively through four stages: immersion, detailed transcription, in-depth episode analysis, and comparative analysis. These stages enabled examination of material, embodied, social, and contextual aspects of challenge-related interactions as they unfolded during rehabilitation.

**Results:** Twenty individuals, comprising four patient–therapist dyads and four triads participated. Challenge was operationalized through three intersecting practices: *(i) Structuring challenge-related work through temporal phases* determined when challenge was introduced, experienced, and consolidated across the session and within tasks; *(ii) Establishing the macro-architecture of challenge* guided what and how it was operationalized by constructing a challenge plan, shaping an optimal bandwidth by tailoring challenge to the person’s abilities and/or experiences, and facilitating its transfer beyond the session; *(iii) Modulating challenge through micro-calibrations* involved moment-to-moment adjustments (e.g., adapting physical assistance, communication style, environmental setup) to modulate active practice, navigate differing perspectives, and cultivate an atmosphere conducive to engaging with challenge. These overarching practices were dynamically enacted through a range of explicit and implicit practices.

**Conclusion:** Challenge in stroke rehabilitation is not merely a task parameter; it is operationalized through an interplay of explicit and implicit practices that structure and enable its responsive implementation. Considering challenge through these practices may help therapists and people with stroke co-construct meaningful experiences of challenge, both within and beyond formal stroke rehabilitation sessions.

## BACKGROUND

Stroke remains a leading cause of disability worldwide, leaving many people with a range of motor, sensory, perceptual, cognitive, communication, and psychological impairments that impact their daily activities, participation in meaningful roles, and quality of life (Christie et al., 2025; Clarke & Forster, 2015; Li et al., 2024). The breadth of these impacts necessitates comprehensive rehabilitation approaches (Christie et al., 2025; Clarke & Forster, 2015; Li et al., 2024). Accordingly, stroke rehabilitation guidelines recommend interdisciplinary interventions, including physiotherapy, occupational therapy, and speech-language therapy, delivered through a combination of functional and impairment-based training, psychosocial support, patient and whānau education, assistive technology, environmental adaptation, and other approaches (Brady et al., 2025; Hildebrand et al., 2023; National Institute for Health and Care Excellence, 2023; Stroke Foundation). These guidelines and interventions are underpinned by core principles (Carey et al., 2019; Khan et al., 2017; Maier et al., 2019). One such principle is ‘challenge’, which is suggested to shape rehabilitation experiences and outcomes (Beaudry et al., 2021; Cherney & van Vuuren, 2022; Maier et al., 2019; Metzger et al., 2014; Signal et al., 2016). Specifically, optimal challenge is thought to promote neuroplasticity and recovery, and foster patient confidence and engagement (Beaudry et al., 2021; Cherney & van Vuuren, 2022; Maier et al., 2019; Metzger et al., 2014; Signal et al., 2016). Consequently, stroke rehabilitation guidelines consistently recommend that rehabilitation tasks are “challenging but achievable” (National Institute for Health and Care Excellence, 2023) and become “progressively more challenging” according to the person’s evolving abilities and goals (Brady et al., 2025; Hildebrand et al., 2023; National Institute for Health and Care Excellence, 2023; Stroke Foundation). For the purposes of this paper, a ‘task’ is understood as any potential catalyst of goal-directed performance within rehabilitation practice, which may operate at an impairment-, activity-, or participatory-level, and be performed as single or multiple repetitions, with or without adaptation. Despite recommendations promoting task challenge, there is limited formalized guidance on how to implement and tailor challenge to the person’s unique needs, preferences, and contexts in clinical practice.

Existing empirical research has focused on relatively simple applications of task challenge that are often unidimensional, easily quantified, and linearly progressed. For instance, substantial evidence supports the use of challenge in motor rehabilitation after stroke, both at the impairment-level through aerobic and strength training (Ammann et al., 2014; Olsen et al., 2023; Ye et al., 2025) and at the activity-level through task-specific training (Gorshkov et al., 2025; Klempel et al., 2023; Woodbury et al., 2016). Emerging research has extended the use of challenge to cognitive and communication rehabilitation through systematic manipulation of task complexity and graded task demands (Gorshkov et al., 2025; Klempel et al., 2023; Tamayo-Serrano et al., 2018). Within these approaches, technology-enabled and home-based programs increasingly facilitate self-directed practice with progressive challenge (Gorshkov et al., 2025; Klempel et al., 2023; Tamayo-Serrano et al., 2018). However, this body of research represents only a fraction of the interventions required to address the diverse needs of people with stroke and is often conducted within highly-controlled, protocol-driven contexts (Chandler et al., 2025; Fu et al., 2025; Li et al., 2024). As a result, existing research provides limited insight into how challenge is operationalized within real-world stroke rehabilitation practice (Chandler et al., 2025; Fu et al., 2025; Li et al., 2024).

Our recent concept analysis study conceptualized challenge as a multifaceted, multidimensional, and dynamic interaction between task demands, the person’s ability, and their subjective experience (Gomes, Alder, Bright, and Signal, 2024). Building on this, our video-reflexive ethnography study found that people with stroke and therapists may co-construct meaningful experiences of challenge through relational processes that integrate lived experience and professional expertise (Gomes, Alder, Bright, and Signal, 2026). These complex, dynamic processes may not be adequately captured within simplified applications or standardized intervention protocols. Instead, viewing challenge as personalized and relationally constructed aligns with growing recognition that stroke rehabilitation requires person-centered approaches supported by collaborative therapeutic relationships to optimize experiences and outcomes (Bishop et al., 2021; Fu et al., 2025; Lawton et al., 2018).

Although current guidelines advocate for holistic, person-centered approaches to stroke rehabilitation more broadly (Brady et al., 2025; Hildebrand et al., 2023; Stroke Foundation, 2025), there is a lack of specific guidance for optimizing challenge (National Institute for Health and Care Excellence, 2023). To help address this gap, this study sought to understand how therapists and people with stroke currently enact challenge in routine clinical practice. Specifically, this study aimed to examine how challenge is operationalized in stroke rehabilitation practice.

## MATERIALS AND METHODS

### Design

This study used a focused video analysis methodology to examine how challenge is operationalized in stroke rehabilitation practice. Focused video analysis leverages researchers’ background knowledge and rich audiovisual recordings to enable detailed analysis of material, embodied, and social interactions as they unfold in real-world contexts (Knoblauch and Schnettler, 2012). Focused video analysis is increasingly used in medical and rehabilitation research as video provides access to communicative, behavioral, and organizational dynamics that may be overlooked through observation or interviews alone (Barthel et al., 2023; Crist et al., 2022; Knoblauch & Schnettler, 2012; Rashid et al., 2019). Given our previous findings that challenge is often dynamic, implicit, and relational in nature (Gomes et al., 2026; Gomes et al., 2024), focused video analysis was well suited to examining how challenge is enacted in clinical practice.

### Ethical approval

Ethical approval was obtained from national, institutional, and organizational committees (HDEC 2023 EXP 13643 and AUTEC 23/196). All participants received a study information sheet and provided written informed consent independently.

### Participants

As this study aimed to examine challenge in routine practice, it required the participation of people with stroke and therapists actively working together in stroke rehabilitation. Accordingly, participants were eligible if they were: (1) an adult with stroke currently working with an eligible therapist in stroke rehabilitation; or (2) a New Zealand-registered physiotherapist, occupational therapist, or speech-language therapist currently working with an eligible person with stroke in stroke rehabilitation. Purposive sampling facilitated diversity in age, gender, ethnicity, and rehabilitation contexts. Additional sampling considerations for people with stroke included time since stroke, and type and severity of stroke-related disability, and for therapists included years of professional experience in stroke rehabilitation. Working patient–therapist dyads and triads were invited to participate through advertisements, research presentations, collaborating networks, and third parties (e.g., colleagues, peers, whānau members).

### Researchers

Focused video analysis encourages researchers to draw on their insider knowledge of both the concept and the field under investigation to accurately interpret participant interactions (Knoblauch, 2001). Such knowledge is not treated as a bias to be eliminated, but as a methodological resource that researchers should reflexively engage with (Knoblauch, 2001). Accordingly, the research team’s positionality was informed by the following backgrounds and expertise: EG is an early-career neurorehabilitation physiotherapist and researcher; GA and NS have extensive clinical and research expertise in physiotherapy and neurorehabilitation; FB has specialist expertise in speech-language therapy and neurorehabilitation, and in video- and practice-based qualitative methodologies. Collectively, the team’s complementary expertise enriched the analysis of challenge-related interactions and supported critical interrogation of assumptions through collaborative discussions (Knoblauch, 2001).

### Data collection

EG collected observational and video data across two routine stroke rehabilitation sessions per patient–therapist dyad or triad. An initial, non-recorded observation session established rapport and contextual understanding. This was followed by a second, video-recorded session using a combination of static and mobile cameras to capture both the broader therapy environment and detailed challenge-related interactions. Observational memos were written during and after each session to document reflexive interpretations and contextual insights.

### Data analysis

Data analysis followed principles of focused video analysis, emphasizing intensive, theoretically informed, and contextually grounded examinations of challenge and its related interactions (Knoblauch & Schnettler, 2012; Knoblauch & Tuma, 2020; Kristensen, 2018). Analysis occurred iteratively through four stages: *immersion, detailed video transcription, in-depth episode analysis,* and *comparative analysis*.

*Immersion* involved the research team repeatedly viewing the video data, with EG immersing in the full dataset. Immersion memos were integrated with observational memos to capture initial impressions of what, when, where, why, and how challenge-related interactions occurred, and who shaped them. *Detailed video transcription* produced extensive transcripts of each video-recorded session, with a particular focus on interactional, relational, temporal, and contextual aspects of challenge. Transcripts embedded timestamps and still images, creating an analytic timeline of each session and supporting traceability to the video data. Descriptive labels were then developed to characterize how challenge was operationalized across each timeline. *In-depth episode analysis* drew on principles of crystallization to enhance data interpretation (Ellingson, 2009). For example, selected video excerpts of particularly rich or unique challenge-related interactions were further explored through the development of narrative accounts, describing the interaction from each participant’s point of view to consider how they may have individually experienced it (Ellingson, 2009). In parallel, analytic maps were created to examine the sequence and relationships between descriptive labels, visualizing how challenge-related interactions unfolded across each timeline. Finally, *comparative analysis* involved constant comparison across video footage, memos, transcriptions, labels, narratives, and maps to test and refine emerging interpretations of how challenge was operationalized in stroke rehabilitation practice.

### Rigor

Rigor was established through strategies for enhancing trustworthiness and observational credibility (Ahmed, 2024; Coker et al., 2013; Knoblauch, 2001). Analysis was grounded in the research team’s conceptual and clinical expertise (Knoblauch, 2001). Analytic processes were iterative, transparent, and reflexive, with immersion and memo writing used to document an audit trail of evolving interpretations (Ahmed, 2024; Rashid et al., 2019). Thick, multilayered representations of the data supported fidelity and confirmability (Ahmed, 2024; Ellingson, 2009; Rashid et al., 2019). Crystallization was used to deepen interpretation, while constant comparison supported coherence across analyses (Ellingson, 2009; Rashid et al., 2019). Frequent research team discussions were used to interrogate assumptions, consider alternative interpretations, and integrate diverse theoretical and clinical perspectives, thereby strengthening credibility (Ahmed, 2024; Rashid et al., 2019).

## RESULTS

In total, 20 participants were included, encompassing four patient–therapist dyads and four triads, involving eight people with stroke, two supporting whānau members, nine therapists, and one therapy assistant. Demographic characteristics of people with stroke and therapists are summarized in Tables 1 and 2, respectively. Participants represented diverse rehabilitation contexts, including across rehabilitation disciplines (i.e., physiotherapy, occupational therapy, speech-language therapy), settings (e.g., outpatient clinic, person’s home), foci (e.g., fatigue management, psychosocial education, cognitive communication, indoor walking), and the duration of the patient–therapist working relationship, as detailed in the Supplementary file. Data collection generated 16 hours of observation and eight hours of video data from participants’ routine stroke rehabilitation sessions, with video sessions ranging from 24–78 minutes.

**Table 1.** Demographic characteristics of people with stroke (n=8)

| **Pseudonym** | **Age** | **Ethnicity** | **Time post-stroke** | **Severity of stroke disability** | **Low-scoring ($\leq 2$) HRQOL domains** | **Type of rehabilitation service used** |
| --- | --- | --- | --- | --- | --- | --- |
| **Clara** | <60 years | Other European | 4–5 years | Slight disability | Hand function | Private outpatient/community |
| **Sylvia** | 65–70 years | NZ European | 2–3 years | Moderate disability | Hand function<br>Mobility<br>ADLs | Private outpatient/community |
| **Eleanor** | 71–80 years | NZ European | 1–3 months | Moderately severe disability | Mobility<br>ADLs | Private outpatient/community |
| **George** | 71–80 years | NZ European | 4–6 months | Slight disability | Social participation<br>Memory and thinking | Public outpatient/community |
| **Helena** | >80 years | Other European | 4–6 months | Moderate disability | Strength<br>Hand function<br>ADLs<br>Social participation | Public outpatient/community |
| **Lillian** | 71–80 years | NZ European | 4–6 months | Moderately severe disability | Hand function<br>Mobility<br>ADLs<br>Communication<br>Mood | Private outpatient/community |
| **Ray** | 60–64 years | NZ European | 1–3 months | Moderate disability | Communication | Public outpatient/community |
| **Ana** | 60–64 years | Māori | <1 month | No significant disability | Social participation | Public outpatient/community |

*Key.* ADLs, activities of daily living; HRQOL, health-related quality of life; NZ, New Zealand; <sup>a</sup>Classified by the therapist using the Modified Rankin Scale (van Swieten et al., 1988); <sup>b</sup>Self-reported on the Short-Form Stroke Impact Scale (MacIsaac et al., 2016).

**Table 2.** Demographic characteristics of rehabilitation therapists (n=9)

| Category | Demographic Characteristic | n |
| --- | --- | --- |
| **Rehabilitation discipline** | Physiotherapy | 5 |
|  | Occupational therapy | 3 |
|  | Speech-language therapy | 1 |
| **Years of professional experience in stroke rehabilitation** | <2 years | 2 |
|  | 2–5 years | 1 |
|  | 6–10 years | 1 |
|  | 11–15 years | 3 |
|  | 16–20 years | 1 |
|  | 20+ years | 1 |
| **Type of rehabilitation service provided** | Private outpatient/community | 5 |
|  | Public outpatient/community | 4 |

Focused video analysis illustrated that challenge in stroke rehabilitation was dynamically operationalized through three overarching practices. *Structuring challenge-related work through temporal phases* described when challenge-related practices occurred across a session and within tasks. Within these phases, *Establishing the macro-architecture of challenge* referred to what and how challenge was operationalized. These two overarching practices commonly involved ‘explicit’ practices that were clearly verbalized, formalized, and/or overtly related to the operationalization of challenge. *Modulating challenge through micro-calibrations* characterized refinement of the phases and macro-architecture of challenge. This overarching practice commonly involved ‘implicit’ practices that were more subtle, responsive, and entailed moment-to-moment adjustments, typically occurring through indirect cues and/or with little to no verbalization, and shaped the operationalization of challenge.

Although these practices are presented distinctly for clarity, they were highly interdependent, reflecting the complex nature of operationalizing challenge in stroke rehabilitation practice. To support interpretation, Figures 1a–d provide a progressive schematic of how each practice may unfold and intersect across a routine stroke rehabilitation session.

**Figure 1a.**
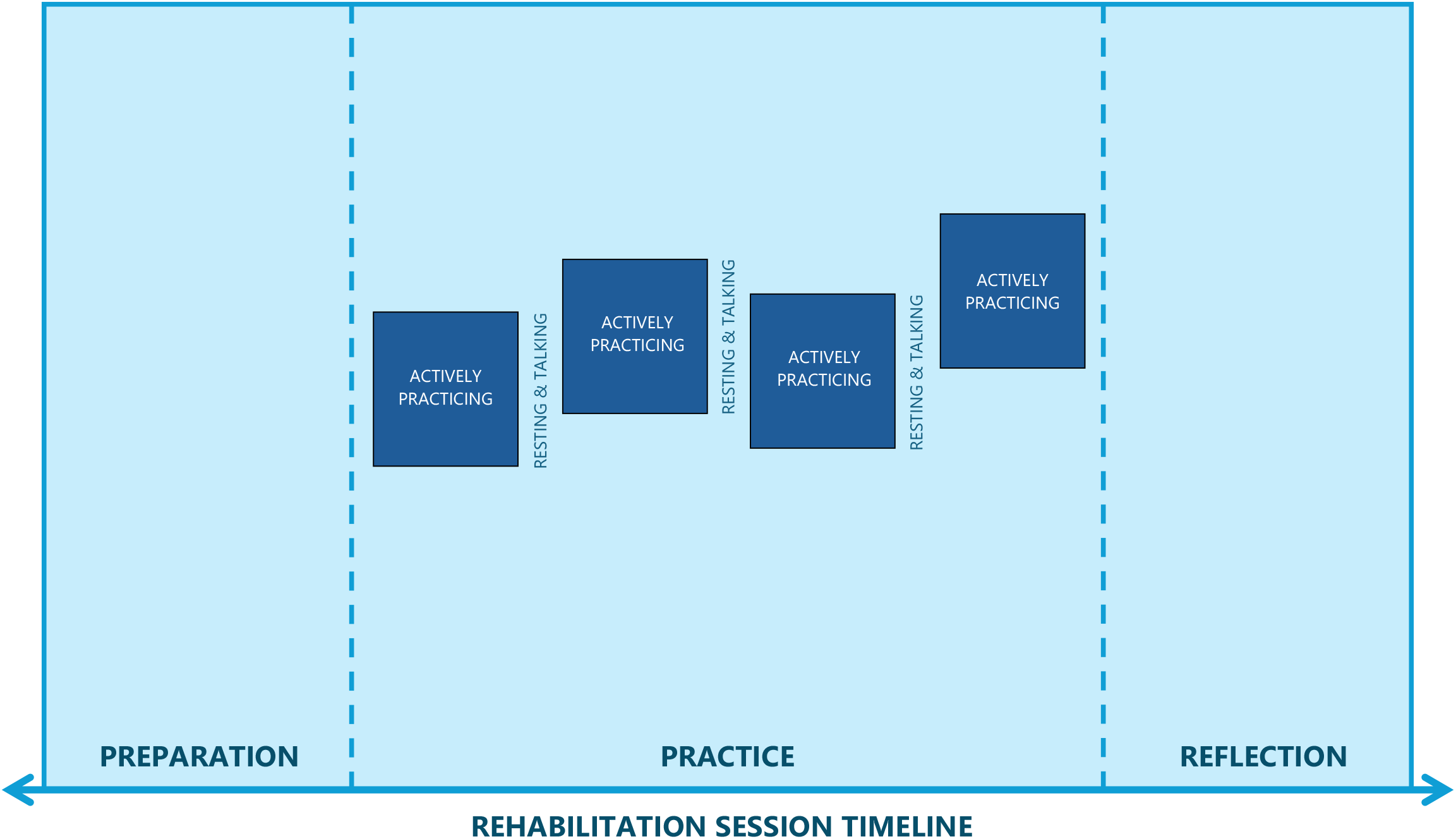
Illustrative example of structuring challenge-related work through temporal phases across a routine stroke rehabilitation session *Note.* Structuring of challenge-related work through temporal phases, which reflects *when* different challenge-related work occurred across a session and within tasks, is represented by shades of blue. The horizontal arrow represents challenge-related work unfolding across the full rehabilitation session timeline. Vertical dashed lines divide the three temporal phases across the session: **Preparation**, **Practice**, and **Reflection**. Within the Practice phase, the colored boxes represent the recurring subphases of **Actively practicing challenge**; each box represents a task, which may involve practicing the same task repeatedly or practicing different tasks. Variations in vertical positioning symbolize varying levels of task challenge. Gaps between the colored boxes represent the recurring subphases of **Resting from challenge** and **Talking about challenge** between tasks.

**Figure 1b.**
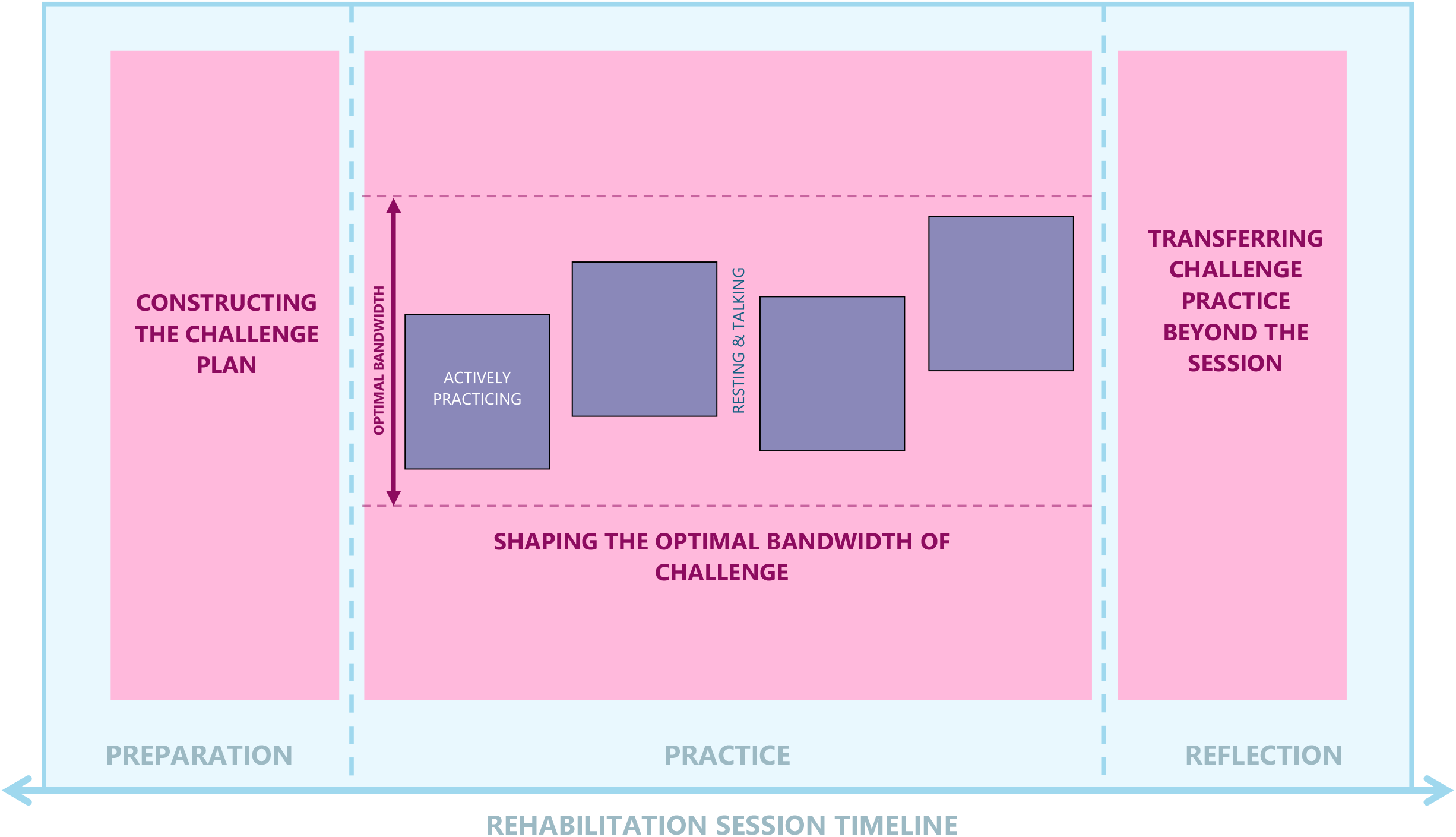
Illustrative example of establishing the macro-architecture of challenge across a routine stroke rehabilitation session *Note.* Establishing the macro-architecture of challenge, which reflects *what* and *how* task challenges were selected, tailored to the person, and contextualized to broader rehabilitation goals, is represented in pink. Within the Preparation phase, the pink area represents when acts related to **Constructing the challenge plan** typically occur. Within the Practice phase, the pink area represents when acts related to **Shaping the optimal bandwidth of challenge** typically occur; the double-pointed vertical arrow illustrates the range of challenge levels within the bandwidth, and the horizontal dashed lines illustrates the upper and lower boundaries of the bandwidth. Within the Reflection phase, the pink area represents when acts related to **Transferring challenge practice beyond the session** typically occur.

**Figure 1c.**
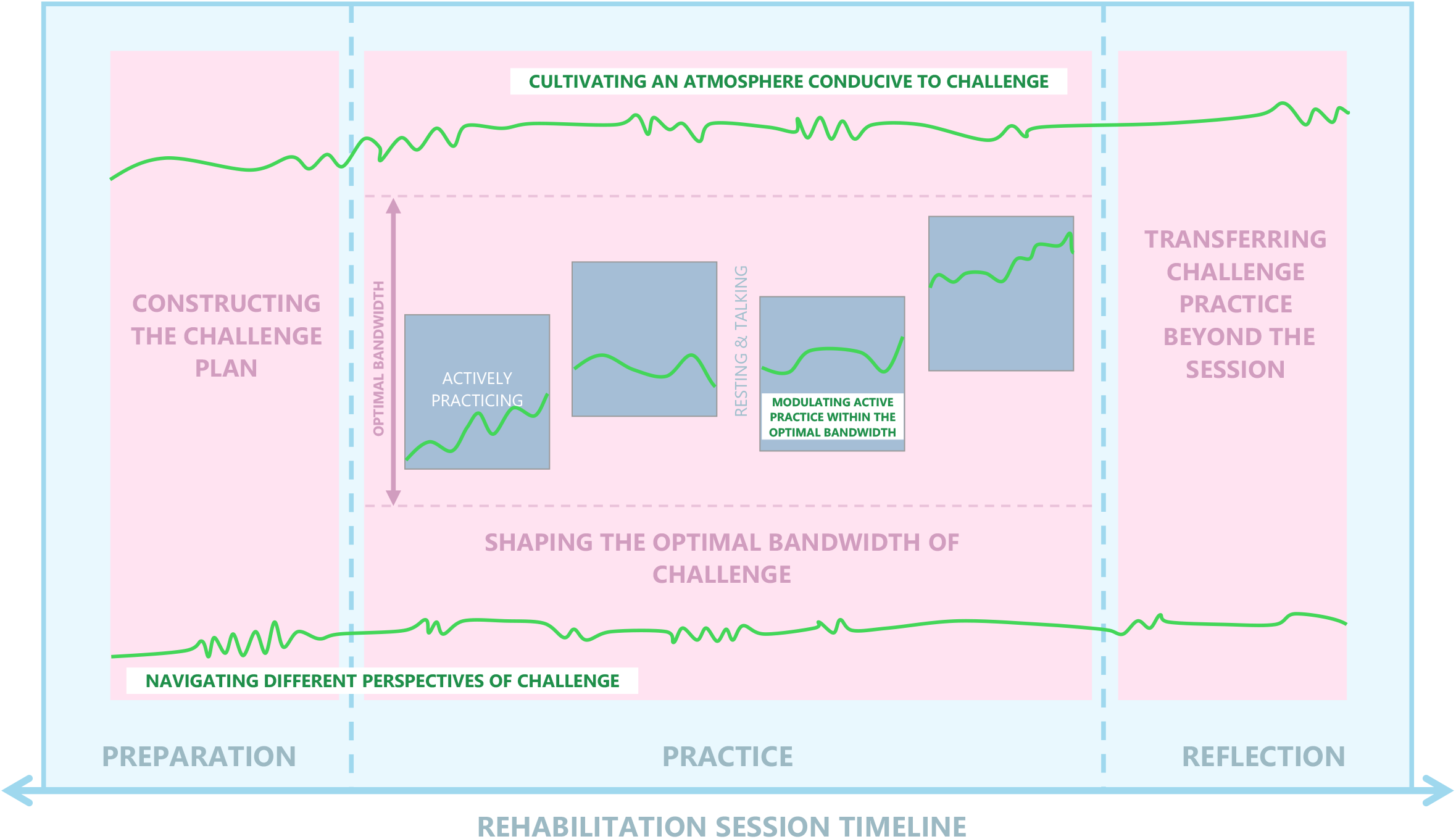
Illustrative example of modulating challenge through micro-calibrations across a routine stroke rehabilitation session *Note.* Modulating challenge through micro-calibrations, which reflects the subtle, moment-to-moment adjustments used to fine-tune challenge in response to the person’s dynamic performance, experience, and the therapeutic relationship, is represented in green. Fluctuations in the green lines represent moment-to-moment micro-calibrations. Most prominently fluctuating within the Practice phase and at the end of the session, the upper green line represents when acts related to **Cultivating an atmosphere conducive to challenge** typically occur. Most prominently fluctuating within the Actively practicing challenge subphases, the middle green lines inside the colored boxes represent when acts related to **Modulating active practice of challenge within the optimal bandwidth** typically occur. Most prominently fluctuating during Constructing the challenge plan, and between periods of Actively practicing challenge, the lower green line represents when acts related to **Navigating different perspectives of challenge** typically occur.

**Figure 1d.**
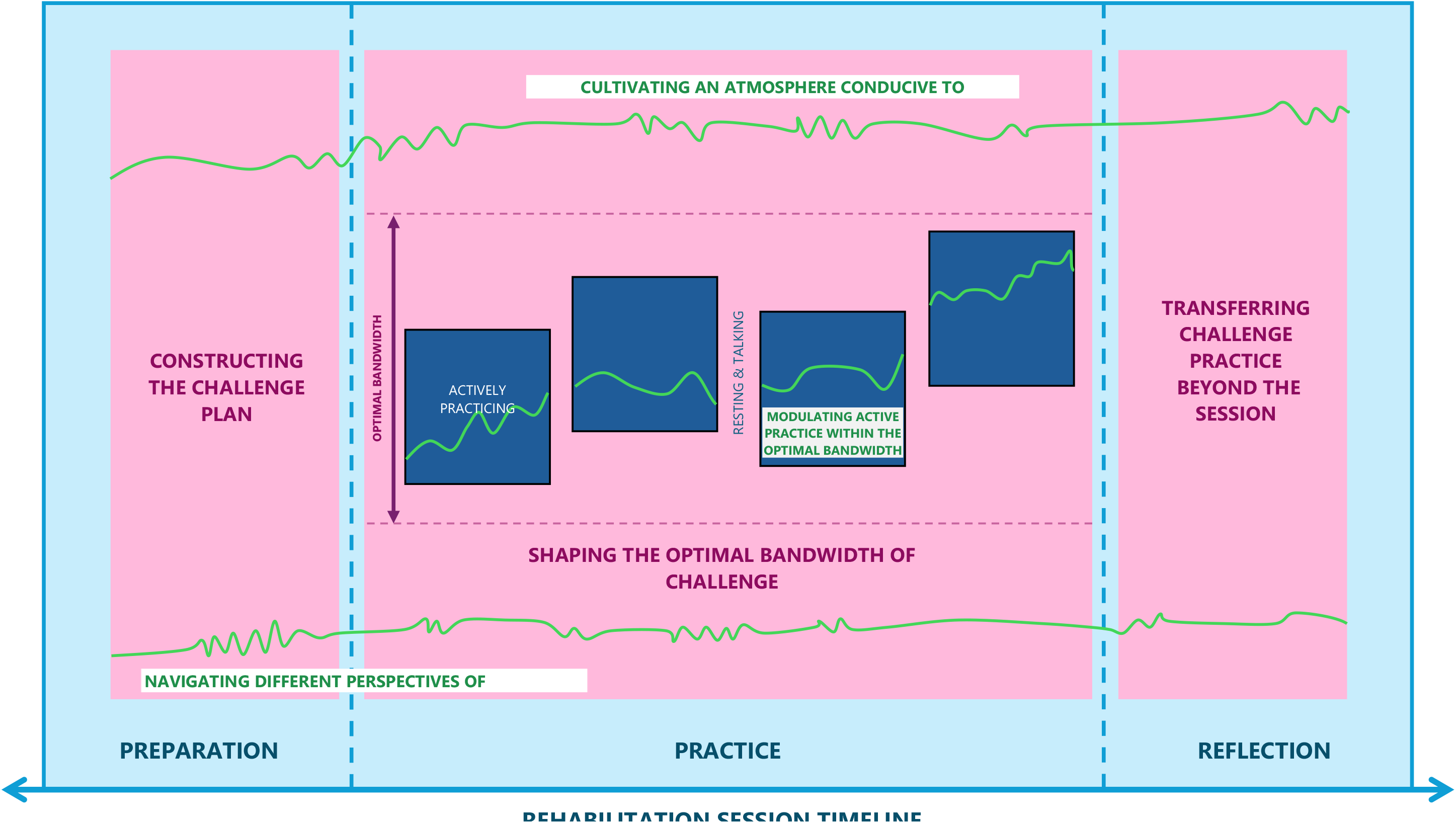
Integrated illustrative example of the three operational practices of challenge in stroke rehabilitation practice *Note.* This integrated figure illustrates how the temporal phases, macro-architecture, and micro-calibrations intersect to shape when, what, and how challenge is operationalized in stroke rehabilitation practice.

### Structuring challenge-related work through temporal phases

Across dyads and triads, challenge-related work unfolded in a similar sequence (illustrated in Figure 1a). These phases followed a rhythm that guided *when* key practices related to operationalizing challenge typically occurred, reflected at two temporal scales: 1) ‘Phases of challenge across the session’ and 2) ‘Subphases of challenge within and between tasks’.

Rather than being confined to individual tasks, the identified ‘Phases of challenge across the session’ reflect when different types of challenge-related work unfolded across the session. Specifically, three phases were identified: 1) Preparation phase, 2) Practice phase, and 3) Reflection phase. These phases created a temporal structure for when challenge was explicitly introduced, experienced, and consolidated.

First, the Preparation phase typically occurred early in the session, following a brief exchange of pleasantries and rapport-building, after which people with stroke and therapists turned their attention to explicitly considering challenge.

> *For example, while seated together in the clinic, a physiotherapist initiated with an open prompt of “How’s your week been?” The person responded with an upbeat “Good!”, before steering the conversation toward how they had been managing challenges related to walking since their last session. The person described practicing walking to their mailbox but struggling with fatigue when navigating their driveway, and expressed worry about managing it alone when their spouse travels next week.*

Such conversations surfaced the person’s current challenges, and at times, informed subsequent planning of challenge practice.

<u>Second, the Practice phase typically occurred in the</u> middle of the session and was characterized by engaging with task challenge(s). This was often marked by a visible shift to a task-specific setting or use of task-specific materials.

> *Continuing with the previous example, following their seated discussion, the person and physiotherapist moved to the treadmill. During treadmill walking, challenge became increasingly evident by the physiotherapist gradually progressing the walking speed, explicitly encouraging a longer step length and forward eye gaze, and the person themselves reducing physical support by removing their hand from the rail. This occurred over several one-minute rounds, interspersed with comments from the person about the task becoming “tiring” and their legs “feeling wobbly”, and from the therapist encouraging them to “keep pushing” and acknowledging they were “working really hard”, indicating effortful challenge.*

Third, the Reflection phase typically occurred toward the end of the session, where challenge was consolidated and contextualized through reflective dialogue.

> *For example, after completing several task challenges, including treadmill walking, standing balance tasks, overground walking, and tennis drills, the person and physiotherapist returned to their seats to debrief about how the person now felt about walking to their mailbox alone.*

Shared understanding of the phases appeared to support anticipation and smoother transitions between phases for both people with stroke and therapists, particularly in instances where challenge seemed optimal. However, the rhythm of challenge-related work was also influenced by the session duration, which constrained how much time and priority was given to each phase.

> *For example, in a different session that was shortened due to a delayed start, the physiotherapist and occupational therapist prioritized practicing challenge from the beginning to the end of the session, and apologized to the person for the lack of dedicated preparation and reflection time.*

This suggested that, while the more conversational phases of challenge-related work were valued, therapists prioritized the practice phase when time was limited.

Within the middle, Practice phase of the session, the ‘Subphases of challenge within and between tasks’ reflected when different types of challenge-related occurred during task practice. Specifically, three subphases were identified: 1) Actively practicing challenge, 2) Resting from challenge, and 3) Talking about challenge.

First, during the Actively practicing challenge subphase, the therapist typically facilitated the person’s direct engagement with task challenge, which was often progressively adapted over time.

> *For example, an occupational therapist initially set up an easier task of grasping a large, lightweight, empty cup from the dining table to explore the person’s ability. As the person demonstrated greater control and appeared more at ease, the occupational therapist progressed the task challenge to grasping a smaller, heavier cup filled with water, requiring greater precision and strength.*

Such practices appeared to represent the core therapeutic work of challenge, which was central to promoting functional improvement and enhancing involvement in meaningful activities.

Second, the Resting from challenge subphase typically involved breaks which enabled the person to gain physical, cognitive, and emotional respite between tasks. The duration and frequency of rests varied based on the needs of the person and the nature of the task.

> *Continuing with the previous example, in the occupational therapy context, as the person completed more repetitions of the cup-grasping task, upper limb fatigue increased and required longer rests before re-engagement. In contrast, in a speech-language therapy context, as a person became more confident with using a word-finding strategy, they needed fewer breaks.*

These examples illustrate how rest appeared integral to pacing active practice of challenge across both the task and the session.

Third, Talking about challenge typically involved in-depth conversations which often occurred during rests. Talking appeared to provide an opportunity for therapists to ‘check in’ and connect with the person in between periods of active practice. However, the content of these conversations varied.

> *For example, early in the practice of the cup-grasping task, the person’s and occupational therapist’s conversations often focused on validating effort, gaining patient-reported challenge ratings, providing feedback, and adapting the task to optimize challenge for the next attempt. As the task became better optimized, these conversations centered less around task challenge and became increasingly casual and relational. For instance, later in the practice, the occupational therapist instead spent this time briefly encouraging the person’s success before sharing a light personal story about their cat—a shared interest—while the person rested.*

Such personal conversations appeared to support deeper therapeutic connection while temporarily easing the intensity of challenge-related work, before active practice resumed.

Therapists tended to lead the active practice and talking subphases, maintaining control over the delivery and narrative of challenge. In contrast, people with stroke tended to lead the resting subphase, using it to regulate their capacity to sustain active practice. This pattern seemed to reflect each participant’s perceived expertise and responsibilities, and shaped when therapists and people with stroke exercised power and agency over challenge-related work. Furthermore, when a shared understanding of the rhythm of challenge-related work was established, this allowed therapists to encourage the person to “give it a go” knowing there would be an opportunity to talk about and adjust challenge if needed, and allowed people with stroke to “work hard” knowing rest would follow. However, when challenge appeared suboptimal, this rhythm seemed to become less predictable, with abrupt shifts in control leading to more reactive transitions between subphases.

> *For example, a physiotherapist appeared to intentionally minimize time for resting and talking about unsuccessful task practice by moving quickly to a different task challenge (e.g., from a cardiovascular to a balance task). This diminished opportunities for the person to critique or express negative experiences of active practice. Similarly, when a physiotherapist expressed concern about a person’s heavy breathing but the person felt able to continue, the person shortened their rest period and independently resumed active practice, asserting their own judgement about challenge.*

Overall, *Structuring challenge-related work through temporal phases* illustrates that challenge is not operationalized solely through active practice of a task, but through a therapeutic rhythm that allows both people with stroke and therapists to anticipate and structure *when* various aspects of challenge-related work occur across the session and within and between tasks.

### Establishing the macro-architecture of challenge

The macro-architecture describes the operational strategies used across the session and within the task phases to guide *how* and *what* task challenges were operationalized (illustrated in Figure 1b). This encompassed three key practices: 1) ‘Constructing the challenge plan’, 2) ‘Shaping the optimal bandwidth of challenge’, and 3) ‘Transferring practice of challenge beyond the session’.

First, ‘Constructing the challenge plan’ typically occurred within the early, Preparation phase of the session, where an explicit plan for subsequent practice of challenge was often constructed. This involved identifying the task challenge(s) that would be targeted through active practice. The extent to which the person and therapist collaborated in this planning process varied.

> *For example, in one dyad, the challenge plan was co-constructed. Guided by the person’s priorities, the person and physiotherapist collaboratively identified that improving the person’s confidence in walking up and down stairs was essential to their independence at home and for attending an upcoming holiday in an unfamiliar environment. In contrast, in another dyad where the person had mild residual impairments, an occupational therapist adopted a more pre-determined approach to constructing the challenge plan. Drawing on the equipment available, the occupational therapist proposed a pinch-grip exercise using tweezers and a cotton ball, explaining how this could support fork manipulation, if the person happened to find it challenging in the future.*

Differences in planning dynamics appeared to be shaped by the rehabilitation setting. In home-based rehabilitation, people with stroke tended to take on the role of identifying what challenges were available and pressing, whereas in clinic-based settings, therapists tended to assume this role.

Many dyads and triads also broadly planned specific task challenge parameters and desired outcomes of active practice. This explicit planning appeared to foster mutual clarity and enhance the person’s physical and emotional readiness for challenge. However, in some instances, these parameters and outcomes were deliberately left open-ended *(e.g., “Let’s see how we go”)*. This was most common when tasks appeared to be highly meaningful and potentially pressurizing, when the person’s capability or readiness was uncertain, or when providing too much information risked overwhelming the person or raising tensions.

> *Continuing with the previous example, when the person first attempted the stairs, they experienced significant fear and required hands-on assistance. The physiotherapist normalized this response as “part of the process” and suggested revisiting independent practice later. However, toward the end of the practice phase, once the person had experienced repeated success with minimal assistance, the physiotherapist highlighted this improvement as clear evidence of progress, while acknowledging that independent stair use was unlikely for now.*

This example illustrates the benefits of combining co-constructed and open-ended challenge plans. However, when plans remained entirely open-ended, they risked obscuring how task challenges were related to the person’s priorities and could privilege a therapist-led planning process.

Second, ‘Shaping the optimal bandwidth of challenge’ typically occurred during the middle, Practice phase of the session, where people with stroke and therapists explicitly aimed to shape an ‘optimal bandwidth’ of challenge for active practice. This optimal bandwidth encompassed a range of task challenges that were considered acceptable based on the person’s dynamic abilities and experiences throughout practice, rather than a singular optimal challenge level or version of a task. To foster early success and build momentum, therapists appeared to initially lead this process by ensuring the person started with familiar and likely achievable versions of the task. This helped shape the easier, lower boundary of the optimal bandwidth of challenge for that task.

> *For example, when first walking down the stairs, where the person perceived high levels of challenge due to fear (evidenced by indicators such as hesitant stepping and high patient-reported challenge ratings), the physiotherapist provided continuous and substantial hands-on assistance to facilitate both physical and emotional safety.*

In contrast, the harder, upper boundary of the optimal bandwidth was often developed through trial-and-error or avoided.

Once the task’s optimal bandwidth of challenge was provisionally outlined, its boundaries were dynamically adapted. In most dyads and triads, this involved gradually progressing towards harder versions of the task and/or narrowing the boundaries of the bandwidth.

> *Continuing with the previous example, once the person’s fear during stair practice began to subside, the person and physiotherapist agreed to incrementally progress the level of physical and emotional challenge. This was achieved by having the physiotherapist gradually withdraw their hands-on assistance, but remain hovering close by to provide minimal assistance if needed.*

This illustrated how physical and emotional dimensions of a task challenge could be layered within the optimal bandwidth.

> *When the physiotherapist later stepped back to facilitate independence without explicitly discussing this with the person, challenge appeared to exceed the upper boundary of the person’s bandwidth and surpass their acceptable limit of fear. This then led the person to become visibly uncomfortable, resist continuing, and prompted explicit re-definition of the challenge bandwidth boundaries.*

Together, these examples display how alignment and misalignment between the person’s and therapist’s perspectives on what constitutes an optimal bandwidth of challenge could directly influence active practice.

Third, ‘Transferring practice of challenge beyond the session’ typically occurred toward the end of the Practice phase and throughout the Reflection phase of the session, where people with stroke and therapists explicitly worked to consolidate and contextualize active practice of challenge to support transfer into the person’s everyday life. In several cases, this transfer process was linked back to the challenge plan to highlight the person’s progress, identify key takeaways, and reinforce continuity by situating practice within a broader trajectory of rehabilitation and recovery.

> *For example, as the person remained somewhat fearful toward the end of stair practice, active practice concluded by returning to easier versions of the task. Here, the physiotherapist invited the person’s spouse to join the session and provide continuous hands-on assistance, effectively replacing the physiotherapist, while the person practiced the stairs one final time. This provided a real-world example of how challenge could be engaged with beyond the rehabilitation session.*

However, when a meaningful challenge could not be identified during planning or when an optimal bandwidth had not been established, therapists seemed to explicitly support transfer in different ways. This included rationalizing the factors that made challenge practice difficult, identifying aspects that could be approached differently in future sessions, and suggesting strategies for temporarily managing (or avoiding) the challenge within everyday life. These discussions aimed to validate engagement with challenge as a confronting and uncertain, yet still valuable, part of the rehabilitation process.

Overall, *Establishing the macro-architecture of challenge* illustrates the complexity of *what* and *how* challenge is bookended and practiced, and highlights how collaboration within this process can influence whether challenge is optimized or not.

### Modulating challenge through micro-calibrations

People with stroke and therapists used micro-calibrations (i.e., a range of subtle, moment-to-moment adjustments) to implicitly modulate challenge in response to the person’s dynamic performance and experience, and the needs of the therapeutic relationship (illustrated in Figure 1c). This encompassed three key practices: 1) ‘Modulating active practice of challenge within the optimal bandwidth’, 2) ‘Navigating different perspectives of challenge’, and 3) ‘Cultivating an atmosphere conducive to challenge’.

First, ‘Modulating active practice of challenge within the optimal bandwidth’ reflected the use of micro-calibrations to performance to respond to fluctuations in the person’s ability and/or experience of challenge, and in turn, maintain Active practice within the optimal bandwidth. Both therapists and people with stroke appeared to make these adjustments through and in response to subtle verbal and non-verbal cues, including body language, facial expressions, performance quality (e.g., accuracy, control, repeated success, compensatory strategies, effort, confidence), physical and spatial positioning, use of equipment, and transient assistance or feedback. These implicit micro-calibrations were generally accepted as supportive or productive by the other party, provided the overall macro-architecture of challenge remained similar to what had been explicitly discussed.

> *For example, when supporting a person to practice a challenging verbal presentation task, the speech-language therapist moved fluidly between micro-calibrations of feedback, including: withholding feedback (to allow space for problem-solving, momentum, frustration tolerance, and independence); offering brief feedback or encouragement (to facilitate performance without disrupting concentration or practice); and judging when these implicit micro-calibrations were no longer sufficient for maintaining optimal challenge due to repeated errors, prolonged pauses, and visible frustration, and explicit input was required.*

Similar micro-calibrations were observed in physiotherapy and occupational therapy contexts in response to fluctuations in movement performance, where therapists subtly adjusted the timing, type, and amount of facilitation to support independence during active practice.

Escalating from implicit micro-calibrations to more explicit input was typically reserved for situations where there were physical or emotional safety concerns, or engagement was rapidly deteriorating.

> *Continuing from the previous example, in moments where the person wished to persist with the word-finding task independently, they proactively declined the speech-language therapist’s support by thinking-aloud, quickly substituting the target word with an alternative word, or quietly stating “I’ll come back [to it]” before continuing their sentence. However, when the person consistently struggled or became exceedingly frustrated with particular words, they let out a sigh and made eye contact with the speech-language therapist as a signal for support.*

These examples reflect the seemingly strategic use of implicit micro-calibrations to enhance active practice of challenge, managing therapist support so that it helped rather than hindered the person’s engagement and autonomy.

Second, ‘Navigating different perspectives of challenge’ involved using micro-calibrations in communication to manage differences between the person’s and the therapist’s perspectives of challenge. Subtle techniques—such as the use of humor, metaphors, ‘floating’ ideas, guided discovery, reflective pauses, and shifts in eye gaze, facial expression, and body language—allowed both parties to surface and work through such differences in low-risk ways.

> *For example, when a person wanted to walk outdoors unaided but their physiotherapist insisted on the use of a low walking frame to manage fatigue, the person attempted to navigate this difference incrementally. They first acknowledged a point of agreement (“I did feel tired after my walk last week …”), then tentatively introduced a different perspective (“… but we’re not going as far, so I’m just not sure I need [the low walking frame]”), before ultimately asserting their stance through action (*begins walking without aid*), which the physiotherapist accepted.*

Such implicit micro-calibrations during challenge-related negotiations appeared to preventpremature assumptions about the other’s perspective and support the development of shared understanding. This reduced the likelihood that differences would escalate into polarizing or disruptive disagreements.

Whether differences in perspectives of challenge could be managed through implicit micro-calibrations in communication or required more explicit discussion appeared to depend on the importance of aligning perspectives within the broader context of the task.

> *For example, during a standing balance task that involved bouncing a basketball, where the person frequently lost balance yet described the task as “easy”, a physiotherapist responded through micro-calibrations—saying “Oh really? Okay!”, using a brief reflective pause, and subtly standing closer and placing a chair behind the person—to signal a different perspective without explicitly disagreeing. It appeared that, because the task was clinic-based, used the therapist’s equipment, and was unlikely to be practiced outside of the therapist’s supervision, the physiotherapist seemed to judge that micro-calibrations would be sufficient to support physical safety without deterring the person’s confidence. In contrast, when the physiotherapist observed inconsistent and unsafe performance during a repeated sit-to-stand task—including repeated loss of balance, significant reliance on upper-limb support, and poor control when lowering back to sitting—they instead initiated an explicit discussion. As sit-to-stand was central to the person’s daily routine and home exercise program, this discussion appeared necessary to align perspectives on the person’s performance and potential risks, and to negotiate how the task would be practiced safely beyond rehabilitation.*

However, these micro-calibrations could also delay or prevent shared understanding and co-construction of optimal challenge. In some cases, even when the macro-architecture of challenge was explicitly co-constructed, therapists still used implicit micro-calibrations to assert their own perspective of challenge.

> *Continuing with the previous example, after agreeing that the person could practice the sit-to-stand task without upper-limb support, the physiotherapist subtly reframed the task immediately before initiating practice by saying “Let’s just start with your hands on the chair for the first few and see.” This seemed to preserve the appearance of shared decision-making, while limiting opportunities for the person to input or disagree.*

Because these implicit assertions were so subtle and fleeting, they were often absorbed into active practice, unless the therapist explicitly invited input or the person explicitly disagreed and voiced their perspective.

Third, ‘Cultivating an atmosphere conducive to challenge’ reflected that the experience of challenge was not solely shaped by the task, but also by the emotional and relational atmosphere in which challenge practice occurred. Generally, a more positive atmosphere—often characterized by sustained engagement, mutual achievement, and enjoyment—was both conducive to and reinforced by co-constructed challenge.

> *For example, therapists appeared more comfortable and likely to offer direct feedback, explain their clinical reasoning, probe the upper, harder boundary of the person’s bandwidth, and openly acknowledge uncertainty or seek the person’s perspective around challenge. Likewise, people with stroke asked questions more freely, expressed their own reasoning or concerns, and contributed to the macro-architecture of challenge.*

This illustrates how more positive atmospheres supported collaborative optimization of challenge.

However, when the atmosphere was fragile, due to factors such as suboptimal challenge or unresolved differences in perspective, both people with stroke and therapists attempted to use micro-calibrations to restore a more positive atmosphere.

> *For example, during an overhead tricep-extension exercise, a person became increasingly frustrated as fatigue caused their arm to drop repeatedly onto their head, letting out a grunt. The occupational therapist responded with a gentle smile and light-hearted humor about the arm “going on strike,” prompting shared laughter. After a brief rest, the occupational therapist praised the person’s improved control in earlier repetitions and redirected the conversation to asking about their grandchildren to diffuse frustration, before resuming active practice.*

In such moments, the priority appeared to shift from practicing challenge to protecting the person’s self-efficacy and trust in their therapist, with the therapist using micro-calibrations to gradually restore an atmosphere capable of tolerating the emotional weight of challenge. However, in some cases, when an atmosphere conducive to challenge could not be implicitly achieved and this was not explicitly addressed, continued reliance on micro-calibrations risked appearing as ‘toxic positivity’.

> *For example, a physiotherapist seemed to unintentionally minimize the person’s visible distress over not completing a stepping task by offering an upbeat “Nearly!” despite high effort. Similarly, in a different session, where a person confidently attempted a hair tying task and shared a satisfied smile despite missing several strands, the occupational therapist seemed to perceive the person to be lacking awareness.*

In these situations, persistent micro-calibrations could perpetuate differing perspectives and prolong practice of suboptimal challenge, risking mutual disengagement with the task and rehabilitation more broadly.

Overall, *Modulating challenge through micro-calibrations* illustrates how people with stroke and therapists use implicit technical and relational micro-calibrations to facilitate optimal practice and experiences of challenge, while also revealing how implicit relational practices can both enable and constrain power and agency in how challenge is operationalized.

To synthesize these findings, Figure 1d brings the three operational practices of challenge in stroke rehabilitation together into an integrated schematic.

## DISCUSSION

This study provides new insights into the overarching practices that shape how challenge is operationalized within routine stroke rehabilitation. Rather than being confined to fixed parameters or isolated moments of physical, cognitive, or emotional effort (Hayward et al., 2021; Page et al., 2012), challenge was operationalized through both explicit and implicit practices across the rehabilitation session. Explicit practices, such as those commonly observed in the temporal phases and macro-architecture, provided an overall framework for structuring and targeting challenge practice, while implicit practices, such as those commonly observed in the micro-calibrations, enabled responsiveness within and around these structures to support effective, safe, and positive experiences of challenge practice. However, relational dynamics shaped how these practices unfolded, and particularly, whose perspectives were prioritized. Together, these findings suggest that operationalizing challenge requires therapists and people with stroke to attend to technical, relational, and reflective aspects of stroke rehabilitation throughout the session, to support the co-construction and responsive practice of optimal challenge. Implications from these findings are summarized in Table 3 to support reflection on how challenge may be operationalized in stroke rehabilitation practice.

**Table 3.**
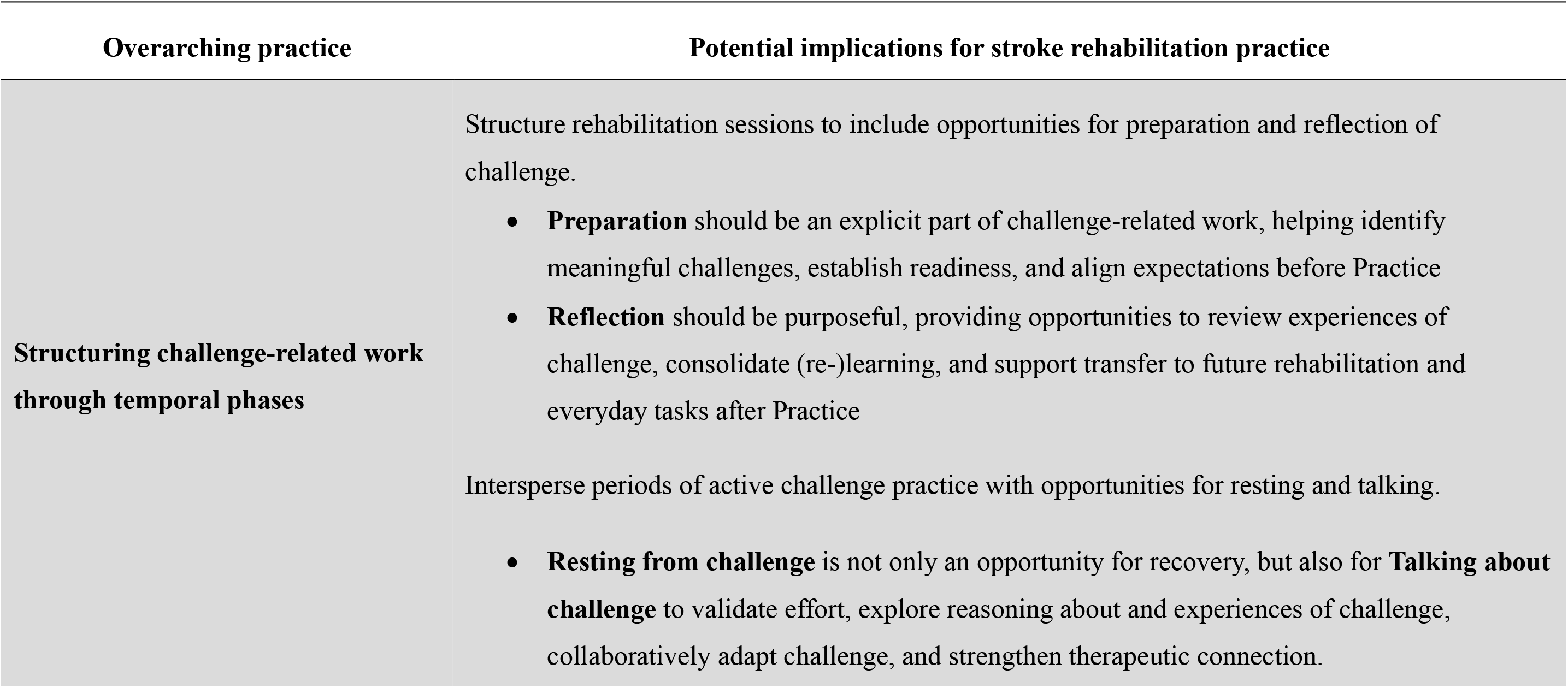
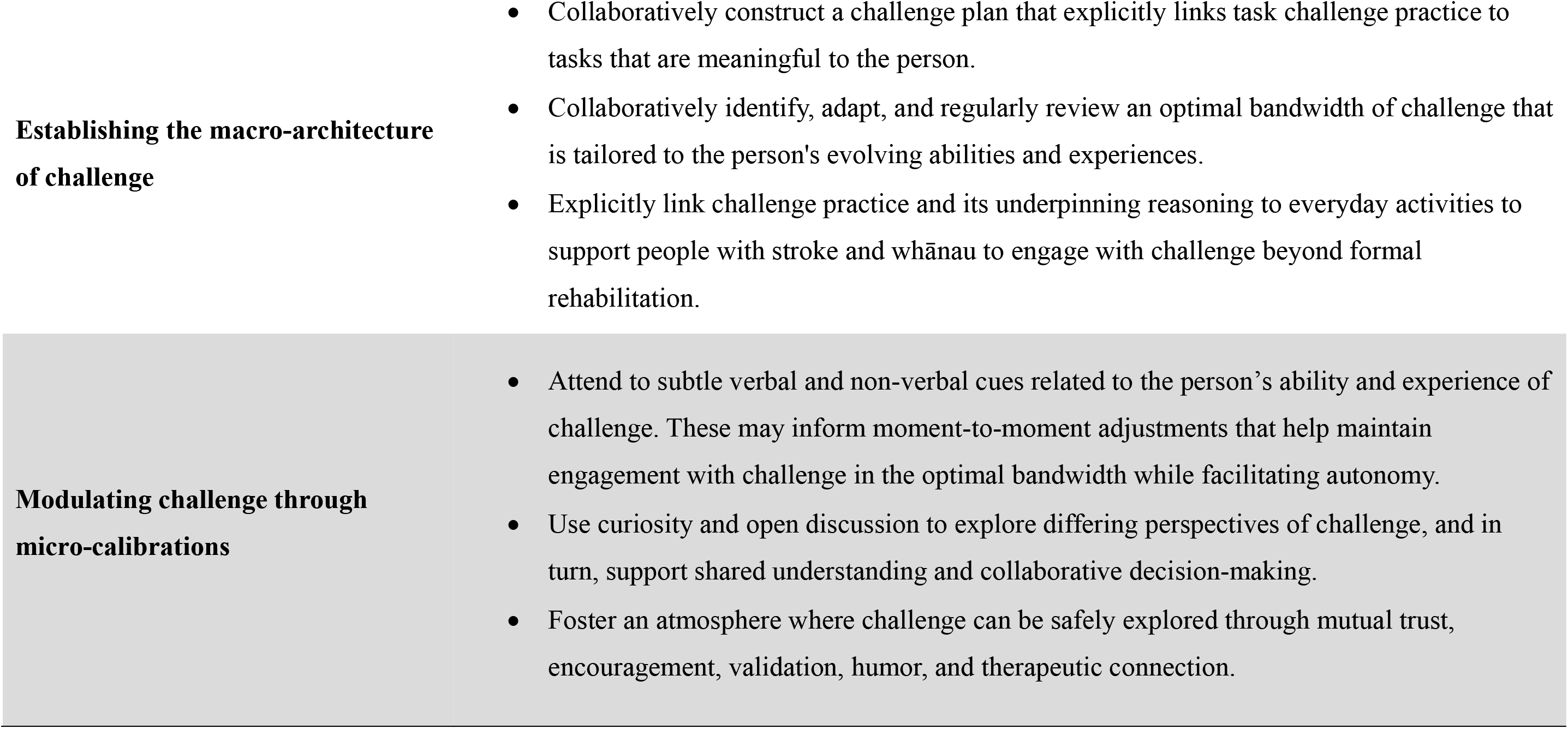
Practice-informed implications for operationalizing challenge in stroke rehabilitation.

### Locating meaningful task challenge

The meaningfulness of the task was a key factor that shaped how the operationalization of challenge unfolded and how the three overarching practices intersected. This extends findings from our earlier interview-based qualitative study (Gomes, Alder, Bright, and Signal, 2026), which found that people with stroke value task challenge when it is aligned with their evolving identity, matched to their perceived capacity at a given point in time, and worthwhile amid competing rehabilitation and everyday life challenges. In this study, ‘Constructing the challenge plan’ around the person’s lived experience helped anchor challenge to meaningful tasks. In turn, this co-construction helped sustain engagement through periods of personal and relational uncertainty. This echoes the shared decision-making literature, which emphasizes collaborative sense-making to determine which of the person’s problems or challenges are important to address (Montori et al., 2023; Tasker et al., 2013; Wottrich et al., 2004). Such processes require therapists to be skilled in guiding conversations to ensure that care is both aligned with the person’s priorities and therapeutically viable (Bishop et al., 2021; Montori et al., 2023).

Indeed, shared decision-making, and developing the authentic working relationship that enables it, are not singular acts but a negotiated continuum that blends both attunement and leadership, and integrates both person- and clinician-centered contributions (Hersh et al., 2017; Kayes & Papadimitriou, 2023; Montori et al., 2023). This highlights the relational work required to enable genuinely collaborative rehabilitation (Hiller et al., 2015). Furthermore, our work showed that locating meaningful task challenges did not only occur during the early Preparation phase where ‘Constructing the challenge plan’ took place. Instead, it was often refined during the middle Practice phase and reinforced during the late Reflection phase of the session, where macro- and micro-adaptations could facilitate connection to challenges of everyday life. These findings counter traditional conceptualizations of task-selection and goal-setting as a discrete and purely cognitive activity that occurs at the outset of rehabilitation, and align with perspectives that portray meaning-making as situational, relational, and emergent (Levack et al., 2015; Sivertsen et al., 2022; Wottrich et al., 2007).

However, our findings also showed that challenge was not always co-constructed. Although therapists were highly skilled at ‘Shaping the optimal bandwidth of challenge’ and ‘Modulating active practice of challenge within the optimal bandwidth’ within tasks, some therapists appeared less confident, equipped, or willing to prioritize alternative tasks when these were more meaningful to the person. This may be compounded by disciplinary norms and organizational priorities, which existing literature suggests may push therapists toward asserting the selection of pre-determined, impairment-focused, and discharge-oriented rehabilitation tasks, which hold variable relevance to people with stroke (Bright et al., 2024; Notkin et al., 2025; Plant et al., 2016).

Together, these findings underscore the value of re-conceptualizing the operationalization of challenge, not as a parameter of a pre-determined task, but as intersecting practices that can be dynamically embedded within different tasks and contexts. In other words, prioritizing the co-construction of a *challenge* plan, rather than a *task* plan, may help people with stroke and therapists to sustain meaningful challenge.

### Relational dynamics of challenge in curating flow experiences

Interactions within our data often reflected the conditions and indicators associated with ‘flow experiences’, particularly when challenge was responsive to the person’s evolving abilities, experiences, and goals (Csikszentmihalyi, 1990). Flow experiences are described as optimal psychophysical states in which individuals are completely immersed in, and intrinsically enjoy, engagement with a task (Csikszentmihalyi, 1990). People with stroke appeared to demonstrate indicators of flow—namely more positive experiences of challenge—when challenges appeared well balanced with the person’s abilities, situated within an environment that supported focused engagement, feedback, and a sense of control, and were linked to meaningful goals. Accordingly, Flow Theory is gaining attention in stroke rehabilitation as a model for using challenge to enhance task engagement and experiences (Lee et al., 2016; Ottiger et al., 2021; Sather et al., 2017; Swanson & Whittinghill, 2015). However, our findings also echo critiques that when a task or goal is highly meaningful, emotional stakes increase, requiring challenge to begin below the person’s ability and be responsively adjusted to maintain emotional safety (Engeser & Rheinberg, 2008; Lawton et al., 2018). This was particularly evident in the acts of ‘Shaping the optimal bandwidth of challenge’ and ‘Cultivating an atmosphere conducive to challenge’, where therapists softened early or tensioned practice of challenge to maintain a positive therapeutic atmosphere. Together, these findings underscore the critical and complex role of therapists in curating positive experiences of challenge (Lawton et al., 2018; Sivertsen et al., 2022; Worrall, 2019).

Reflecting this, emerging extensions of Flow Theory in clinical contexts conceptualize flow as a shared experience between the person and clinician (Pizarro et al., 2020; Riva et al., 2016; Sather et al., 2017). These accounts emphasize the need for clinicians to continuously attune to the person’s unique needs for developing and sustaining flow experiences (Pizarro et al., 2020; Riva et al., 2016; Sather et al., 2017). Similarly, our findings showed that when people with stroke and therapists developed a shared understanding of challenge this often seemed to be underpinned by and further enhance the person’s trust, satisfaction, and enjoyment. Yet, such relational nuances remain largely absent from current applications of Flow Theory in stroke rehabilitation (Lee et al., 2016; Ottiger et al., 2021; Sather et al., 2017; Swanson & Whittinghill, 2015). Collectively, these findings suggest that therapists must continually attune and respond to the person’s experience, to foster positive experiences of challenge.

### Developing a shared artistry of challenge

The nuanced practices identified in this study reflect the ‘professional artistry’ required to optimize challenge in stroke rehabilitation practice. Professional artistry refers to therapists’ dynamic use of explicit and implicit technical, relational, reflective, and creative skills in response to the person and the complexities of clinical practice (Higgs and Titchen, 2001; Paterson, Higgs, and Wilcox, 2005). In this study, artistry was evident in the ways therapists used their clinical expertise to continually respond to the dynamic, sometimes competing, demands of the task, the person’s ability and experience, the therapeutic relationship, and the broader rehabilitation context when operationalizing challenge. However, our findings extend this concept beyond therapists to include people with stroke as active contributors to the artistry required to operationalize challenge (Bright et al., 2017). In particular, people with stroke demonstrated artistry within the acts of Resting from challenge, ‘Modulating active practice of challenge within the optimal bandwidth’, and ‘Navigating different perspectives of challenge’. Within these acts, people with stroke commonly regulated both their own and their therapists’ understandings and practices of challenge, promoting shared understanding and meaningful engagement.

While research suggests patients may develop such artistry implicitly, one way it may be deliberately cultivated is through therapists’ explicit communication of clinical reasoning (Ajjawi & Higgs, 2007; Bishop et al., 2021; Montori et al., 2023). However, the effectiveness of such explication may depend on its timely and context-sensitive delivery, highlighting the importance of therapists’ meta-reasoning in discerning when and how to make reasoning explicit (Ajjawi & Higgs, 2007; Miciak et al., 2018; Sivertsen et al., 2022). Our findings provide insights into the ways in which therapists may deliberately foster the person’s artistry through integrated and dedicated explications of clinical reasoning. Within our data, integrated explications were typically brief and embedded within and between acts of Actively practicing challenge, particularly during Talking about challenge. Here, therapists often reflected strategies described in the literature, such as using informal talk, thinking aloud during decision-making, and disclosing knowledge gaps, to invite the person’s reasoning and explicitly link it to challenge practice (Hersh et al., 2017; Tasker et al., 2013; Thomson, 2010). In contrast, dedicated explications typically occurred during protected, focused time for more comprehensive discussion, such as the early, Preparation and late, Reflection phases of the session. These phases offered structured opportunities for collaborative reasoning about the person’s experiences and management of everyday challenges, aligning with calls for therapists to support people with stroke to navigate challenges beyond the rehabilitation context (Ajjawi & Higgs, 2007; Bishop et al., 2021; Parry, 2009; Sivertsen et al., 2022; Wood et al., 2010).

Overall, supporting people to understand and reason about challenge through the three overarching practices identified in this study may enhance their ability to make sense of and self-direct their engagement with and management of challenge over time ^321, 457^. While previous research has identified practical and social concerns about promoting the person’s artistry, including the time demands and blurring of clinician- and client-centered roles (Hersh et al., 2017; Kayes & Papadimitriou, 2023; Miciak et al., 2018; Wottrich et al., 2007), our findings view the development of the person’s artistry as a valuable therapeutic resource. Indeed, individual and shared artistries may promote shared understanding, streamline the co-construction of meaningful challenge, and support sustained engagement with challenge across the recovery continuum.

### Limitations

This study has several limitations that should be considered when interpreting the findings. While purposive sampling enabled examination of challenge across a range of participants and rehabilitation contexts, inpatient settings, as well as gender and cultural diversity, were underrepresented. As such, the ways in which challenge is enacted in more acute, time-pressured, or culturally diverse rehabilitation contexts may differ from those observed in this study, particularly where opportunities for relational and reflective aspects of challenge may vary. The presence of a researcher and video-recording within rehabilitation sessions may also have influenced participant behavior through a Hawthorne effect; however, such responses may reveal participants’ beliefs about how challenge *should* be operationalized, and therefore, remain insightful (Gwyn, 2002). Additionally, data were collected from only two sessions per dyad or triad, limiting examination of how challenge-related practices evolve over time. Finally, as inherent to video-based qualitative research, analysis is interpretive and shaped by the researchers’ positionality, underpinning theoretical frameworks (Gomes et al., 2026; Gomes et al., 2024), and the analytical process requiring inference of participants’ experiences from observed interactions. While all interpretations were grounded in a range of operational patterns observed within the data, future research could examine the contribution of specific performance-, physiological-, and experience-based measures to challenge-related clinical decision-making. Despite these limitations, this study provides novel, detailed insights into how challenge is operationalized in stroke rehabilitation practice, offering a foundation for researchers and therapists to reflect on how it may be technically and relationally optimized.

## CONCLUSION

This study provides new insights into how challenge is operationalized in routine stroke rehabilitation practice, highlighting an interplay of temporal phases, macro-architecture, and micro-calibrations to shape when, what, and how challenge is practiced and experienced. While the more explicit temporal phases and macro-architecture provided structure to the operationalization of challenge, the more implicit micro-calibrations enabled responsiveness within and around these structures. Together, these findings show that challenge was not contained to a fixed or isolated parameter of a task, but rather, involved continuous, co-constructed practices between the person and therapist that unfolded across the session. Viewing the operationalization of challenge through the overarching practices identified in this study may offer opportunities to develop more meaningful, collaborative, and transferable approaches to challenge and support its application. Collectively, these insights may help optimize the use of challenge within and beyond stroke rehabilitation, and in turn, enhance the experiences and outcomes of people with stroke.

## Data Availability

The dataset generated and analyzed during the study is not publicly available due to its confidential and identifiable nature.

## ACKNOWLEDGEMENTS

We acknowledge the people with stroke, rehabilitation therapists, and support people who shared their time, experiences, and insights for this study. Their contributions were invaluable in shaping both the depth and direction of this work. We also extend our thanks to the organizational leaders who facilitated locality approval and supported recruitment processes.

## DECLARATIONS AND STATEMENTS

- **Funding statement:** This study was supported by a Health Research Council New Zealand Emerging Researcher First Grant (Signal-19/624) and Auckland University of Technology Vice Chancellor’s Doctoral Scholarship.
- **Ethical approval and informed consent statement:** This study was approved by the Northern B Health and Disability Ethics Committee (2023 EXP 13643) on the 28^th^ of July 2023, the Auckland University of Technology Ethics Committee (23/196) on the 28^th^ of August 2023, and by the relevant localities of participating organizations. All participants provided written informed consent prior to enrolment in the study.
- **Disclosure statement:** The authors report there are no competing interests to declare.
- **Data availability statement:** The dataset generated and analyzed during the current study is not publicly available due to its confidential and identifiable nature.

---

**Supplementary file.** Detailed participant demographics and rehabilitation contexts *Key.* ^a^ Classified by the therapist using the Modified Rankin Scale (van Swieten et al., 1988); ^b^ Self-reported on the Short-Form Stroke Impact Scale (MacIsaac et al., 2016).

| Category | Demographic / Context Subcategory | n |
| --- | --- | --- |
| **People with stroke (n=8)** |  |  |
| **Age** | <60 years | 1 |
|  | 60–64 years | 2 |
|  | 65–70 years | 1 |
|  | 71–80 years | 3 |
|  | >80 years | 1 |
| **Gender** | Female | 6 |
|  | Male | 2 |
| **Ethnicity** | New Zealand European | 5 |
|  | Other European | 2 |
|  | Māori | 1 |
| **Phase of stroke** | <1 month | 1 |
|  | 1–3 months | 2 |
|  | 4–6 months | 3 |
|  | 2–3 years | 1 |
|  | 4–5 years | 1 |
| **Severity of stroke disability<sup>a</sup>** | 1: No significant disability | 1 |
|  | 2: Slight disability | 2 |
|  | 3: Moderate disability | 3 |
|  | 4: Moderately severe disability | 2 |
| **All low-scoring (≤2) domains of health-related quality of life<sup>b</sup>** | Strength | 1 |
|  | Hand function | 4 |
|  | Mobility | 3 |
|  | Memory and thinking | 1 |
|  | Activities of daily living | 4 |
|  | Communication | 2 |
|  | Mood | 1 |
|  | Social participation | 3 |
| **Rehabilitation therapists (n=9)** |  |  |
| **Age** | 30–39 years | 3 |
|  | 40–49 years | 3 |
|  | 50–59 years | 3 |
| **Gender** | Female | 9 |
|  | Male | 0 |
| **Ethnicity** | New Zealand European | 3 |
|  | Other European | 3 |
|  | African | 1 |
|  | Indian | 1 |
|  | Australian | 1 |
| **Rehabilitation discipline** | Physiotherapy | 5 |
|  | Occupational therapy | 3 |
|  | Speech-language therapy | 1 |
| **Years of professional experience in stroke rehabilitation** | <2 years | 2 |
|  | 2–5 years | 1 |
|  | 6–10 years | 1 |
|  | 11–15 years | 3 |
|  | 16–20 years | 1 |
|  | 20+ years | 1 |
| **Rehabilitation context of video-recorded sessions (n=8)** |  |  |
| **Rehabilitation setting** | Person's home | 5 |
|  | Outpatient clinic | 1 |
|  | Rest home | 1 |
|  | Hydrotherapy | 1 |
| **Funding model** | Public | 4 |
|  | Private | 4 |
| **All rehabilitation foci (as reported)** | **Symptom management** |  |
|  | Fatigue management | 3 |
|  | Dizziness | 1 |
|  | Emotional lability | 1 |
|  | **Impairment-focused** |  |
|  | Upper limb | 3 |
|  | Sensory | 2 |
|  | Cognitive communication | 1 |
|  | **Activity-focused** |  |
|  | Activities of daily living | 4 |
|  | Balance | 3 |
|  | Indoor walking | 3 |
|  | Outdoor walking | 1 |
|  | Functional communication | 1 |
|  | **Participation-focused** |  |
|  | Sports-related | 2 |
|  | Work-related | 1 |
|  | **Psychosocial support** |  |
|  | Psychosocial education | 3 |
| **Duration of patient–therapist working relationship** | <2 weeks | 1 |
|  | 2–4 weeks | 2 |
|  | 5–8 weeks | 2 |
|  | 9–16 weeks | 2 |
|  | >16 weeks | 1 |

